# Clinical spine care partnerships between low- and high-resource countries: A scoping review

**DOI:** 10.1101/2023.06.07.23291022

**Authors:** Hannah Lin, Kristin Halvorsen, Myat Thu Win, Michael Yancey, Nada Rbil, Abhinaba Chatterjee, Bridget Jivanelli, Sariah Khormaee

## Abstract

**Background:** Clinical collaboration between spine professionals in high-resource (HR) and low-resource (LR) countries may provide improvements in the accessibility, efficacy, and safety of global spine care. Currently, the scope and effectiveness of these collaborations remain unclear. In this review, we describe the literature on the current state of these partnerships to provide a framework for exploring future best practices.

**Methods:** PubMed, Embase, and Cochrane Library were queried for articles on spine-based clinical partnerships between HR and LR countries published between 2000 and March 10, 2023. This search yielded 1528 total publications. After systematic screening, nineteen articles were included in the final review.

**Results:** All published partnerships involved direct clinical care and 13/19 included clinical training of local providers. Most of the published collaborations reviewed involved one of four major global outreach organizations with the majority of sites in Africa. Participants were primarily physicians and physicians-in-training. Only 5/19 studies reported needs assessments prior to starting their partnerships. Articles were split on evaluative focus, with some only evaluating clinical outcomes and some evaluating the nature of the partnership itself.

**Conclusions:** Published studies on spine-focused clinical partnerships between HR and LR countries remain scarce. Those that are published often do not report needs assessments and formal metrics to evaluate the efficacy of such partnerships. Toward improving the quality of spine care globally, we recommend an increase in the quality and quantity of published studies involving clinical collaborations between HR and LR countries, with careful attention to reporting early needs assessments and evaluation strategies.

## Introduction

Musculoskeletal disorders are a leading cause of disability worldwide, with low-resource (LR) countries being the most severely impacted [1, 2]. Spinal disorders and injury have long been recognized as a major public health issue and cause of disability, economic hardship, and morbidity in developed countries. In 2010, the World Health Organization (WHO) Global Burden of Disease study reported that spinal disorders and injuries also place a substantial burden of disability on people in LR countries [3]. This study included both low-income countries (those with a gross national income (GNI) per capita of <$1,085) and lower-middle income countries (those with a GNI per capita between $1,086 and $4,255) as LR countries [4]. In contrast, high-resource (HR) countries have a GNI per capita above $13,206 [4].

LR countries represent 48% of the global population but only 19% of all surgeons, resulting in a ratio of 5.5 providers per 100,000 people compared to 56.9 providers per 100,000 people in HR countries [5]. The surgical specialist workforce is even more inequitably distributed. Major barriers to safe surgical care include limited resources, insufficient surgical workforce, and inadequate training and education programs [6]. Effective partnerships with HR countries provide a potential pathway to addressing some of these challenges.

Historically, some specialized surgical care in LR countries has relied on visiting surgical teams from HR countries to serve selected local patients [6–8]. This model, however, can neglect the importance of investment in local health infrastructure and staff training for more long-term impact [9]. A more sustainable model of high-quality surgical care involves a strong health investment in LR countries, with emphasis on creating sustainable systems with training and resource allocation [6].

In the past few decades, spine-based partnerships involving clinical care and training collaborations between HR and LR countries have arisen as a response to the need for accessible, safe, and affordable spine care globally [10, 11]. Currently, there are several leading organizations, such as World Spine Care (WSC) and the Scoliosis Research Society Global Outreach Program (SRS-GOP), pursuing these clinical spine care partnerships. However, to the authors’ knowledge, there is no centralized summary of all such functioning clinical partnerships.

This scoping review aims to describe the current landscape of peer-reviewed literature reporting on HR-LR country spine-based clinical care partnerships. This will provide a framework for future determination of effective practices and inform the sustainable, equitable, and accountable implementation of future partnerships.

## Methods

### Search Strategy

A curated PubMed search was created using a combination of controlled keywords, including “global health,” “medical missions,” “education,” “training,” “clinical,” “resource limited,” “spine,” and “spinal,” then translated for use in Embase and the Cochrane Library databases. The complete search strategy (S1 Appendix) and a completed PRISMA-ScR checklist (S2 Table) are included for transparency. The search limited publication dates to January 1, 2000 through March 10, 2023, and animal studies were excluded. Covidence, a systematic review software package, was used for deduplication of references, title/abstract screening, full text screening, and data extraction. Reference lists of relevant papers were also screened for potential articles. Each study was screened by three team members to reduce bias.

### Study selection

Three reviewers independently screened titles, abstracts, and full texts obtained from the above search. Articles were included if they were experimental studies, observational studies, or reviews and excluded if they were abstracts or not peer-reviewed. Articles that explicitly reported on partnerships between HR and LR countries or organizations focused on clinical training of healthcare providers and/or direct clinical care of patients were included. All studies reporting on partnerships with an exclusive or significant focus on spine care were included, and studies reporting on partnerships with broader focuses on orthopedic or neurosurgical interventions with no mention of spine-specific interventions were excluded.

### Data extraction and analysis

The following data were extracted from the final included studies utilizing a Covidence-designed standardized extraction form: first author and publication year, reported date range of study partnership, HR country/organization, LR country/organization, reported primary focus of the intervention (eg., spinal trauma, spinal deformity, degenerative/arthritic disease), reported partnership activities (e.g., direct clinical care, clinical training), reported partnership participants (e.g., physicians, nonphysicians), reporting of needs assessments, article’s main focus of evaluation, and main evaluation tools used. Data from studies were independently extracted by two reviewers and discrepancies were resolved by consensus.

## Results

The electronic search retrieved 1528 articles. After removal of duplicate articles, 1271 underwent title and abstract screening and 154 underwent full text review. After additional screening of relevant reference lists, 19 unique papers were included in the final review (Fig 1) [1, 10, 12–28]. Extracted data from all included articles can be found in Table 1.

**Fig 1.**
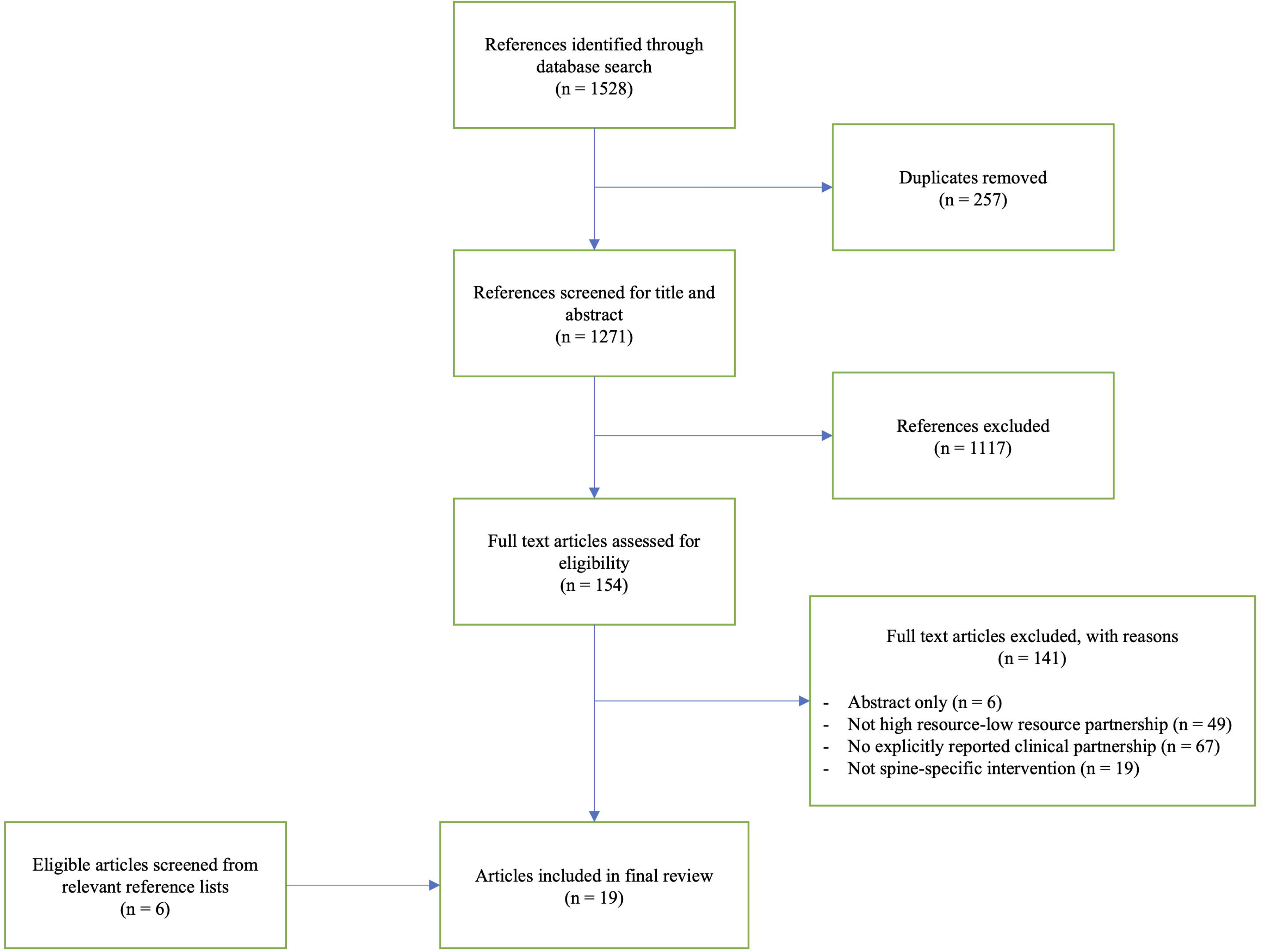
The study selection flowchart.

**Table 1.**
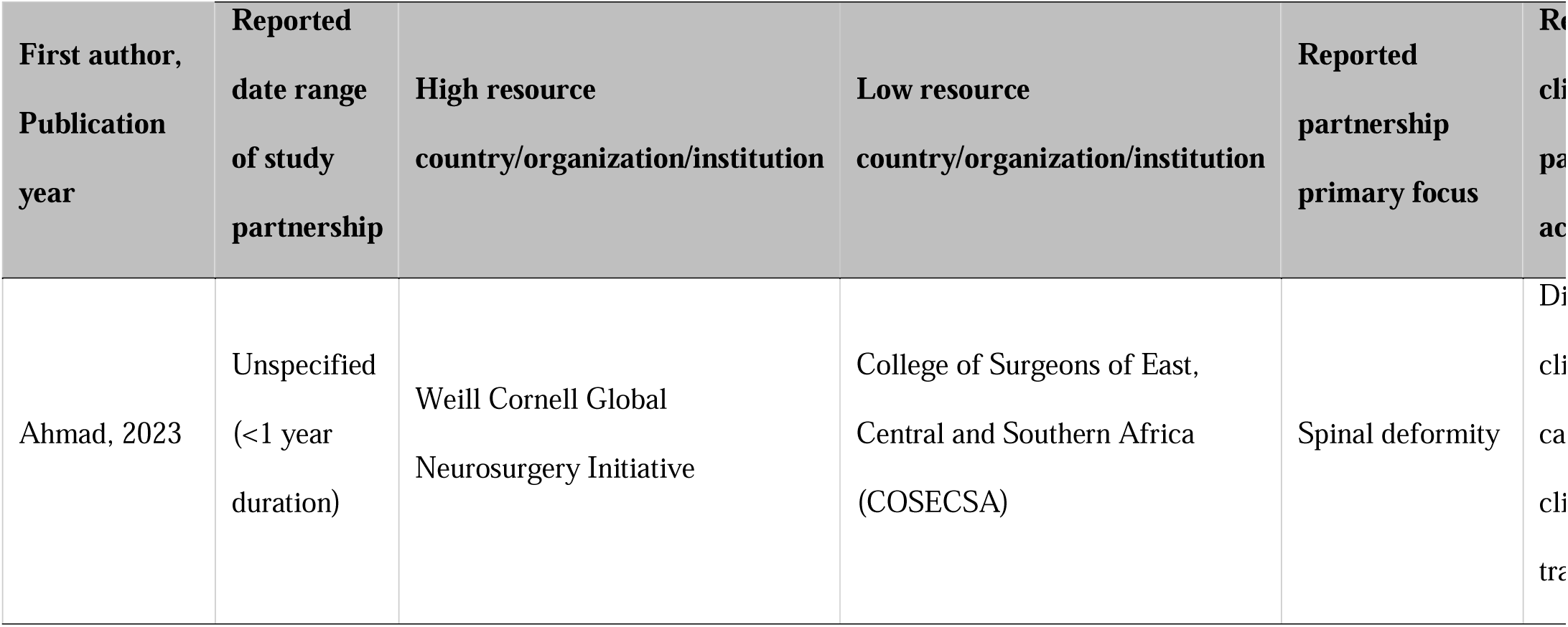
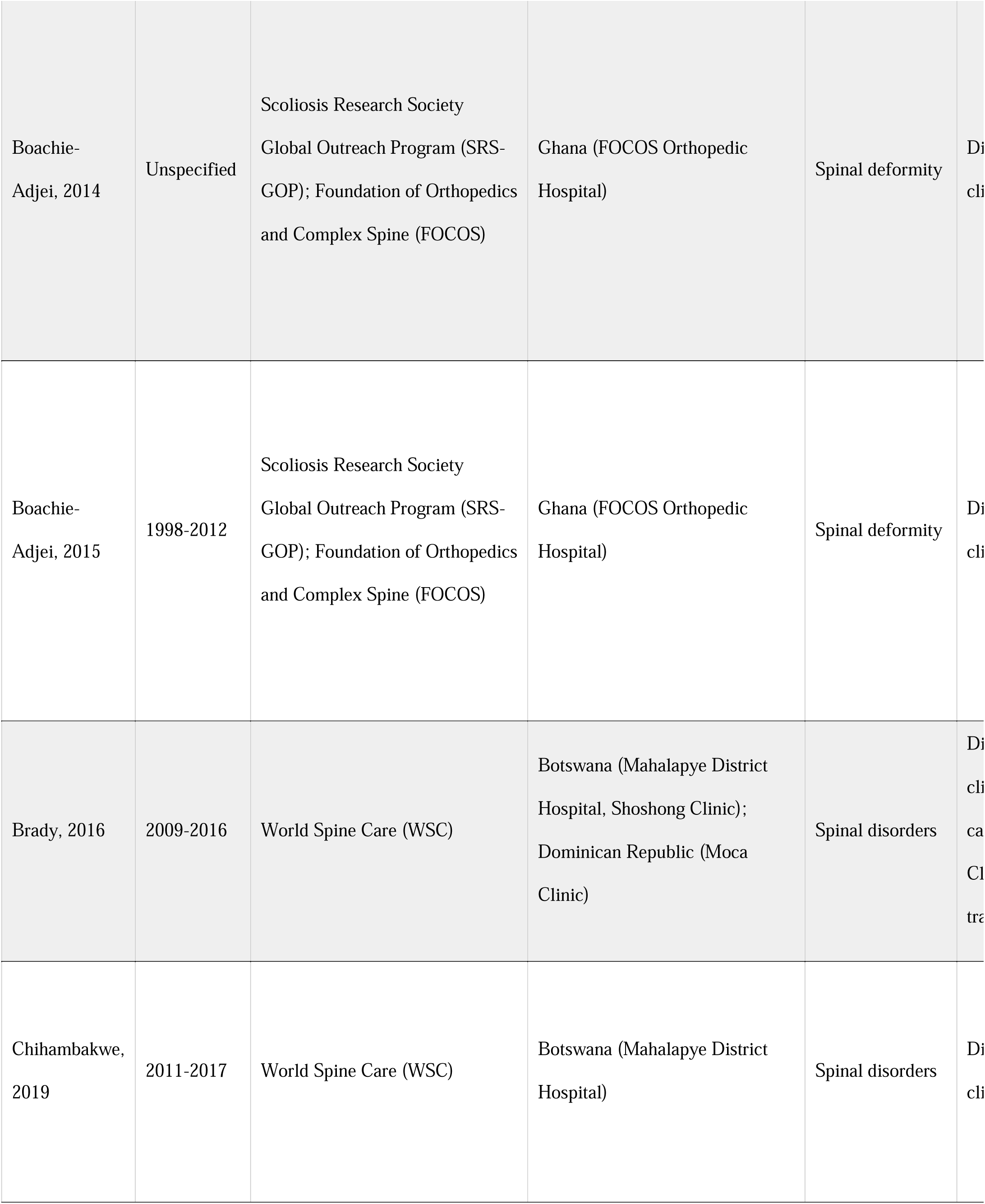
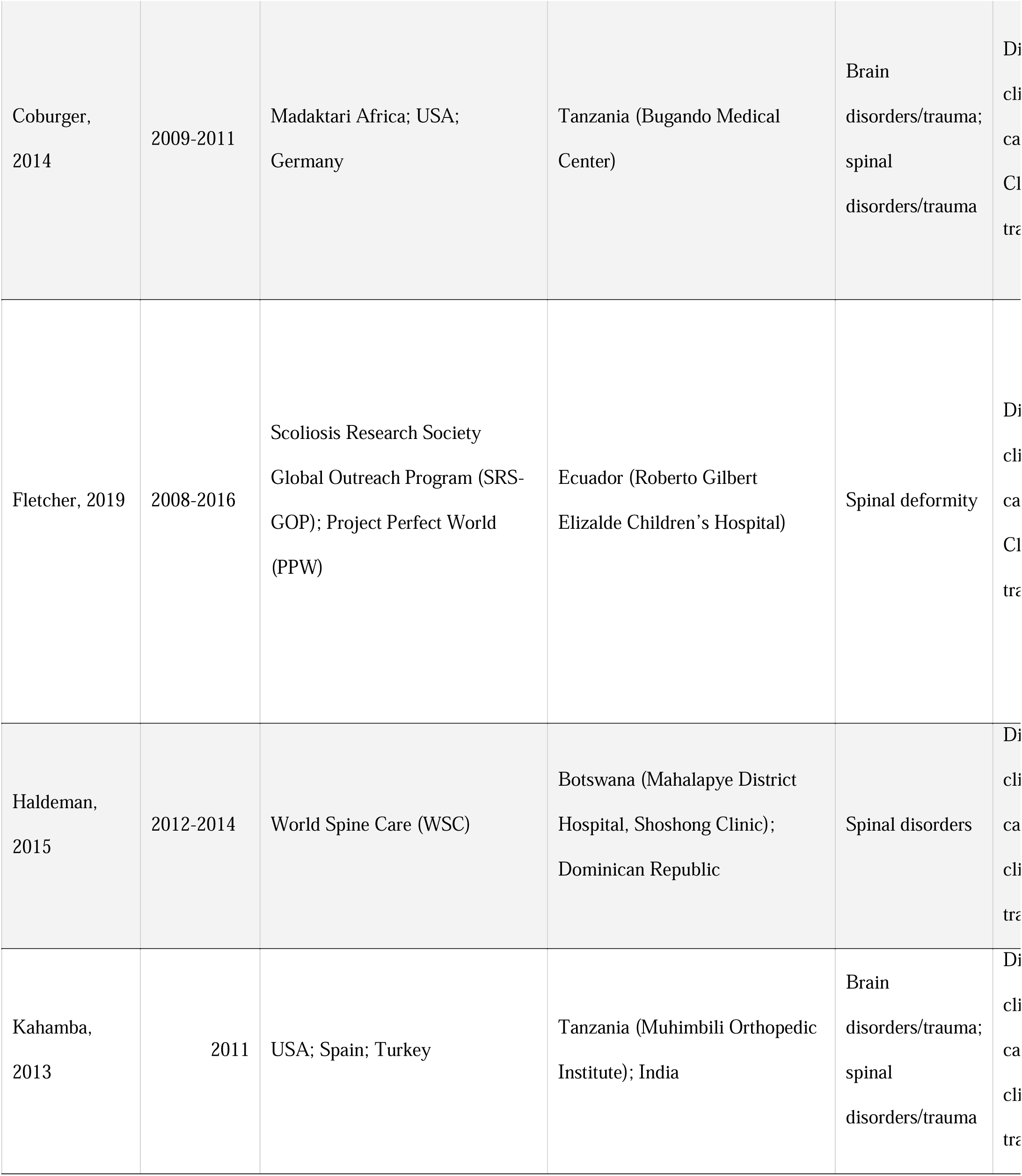
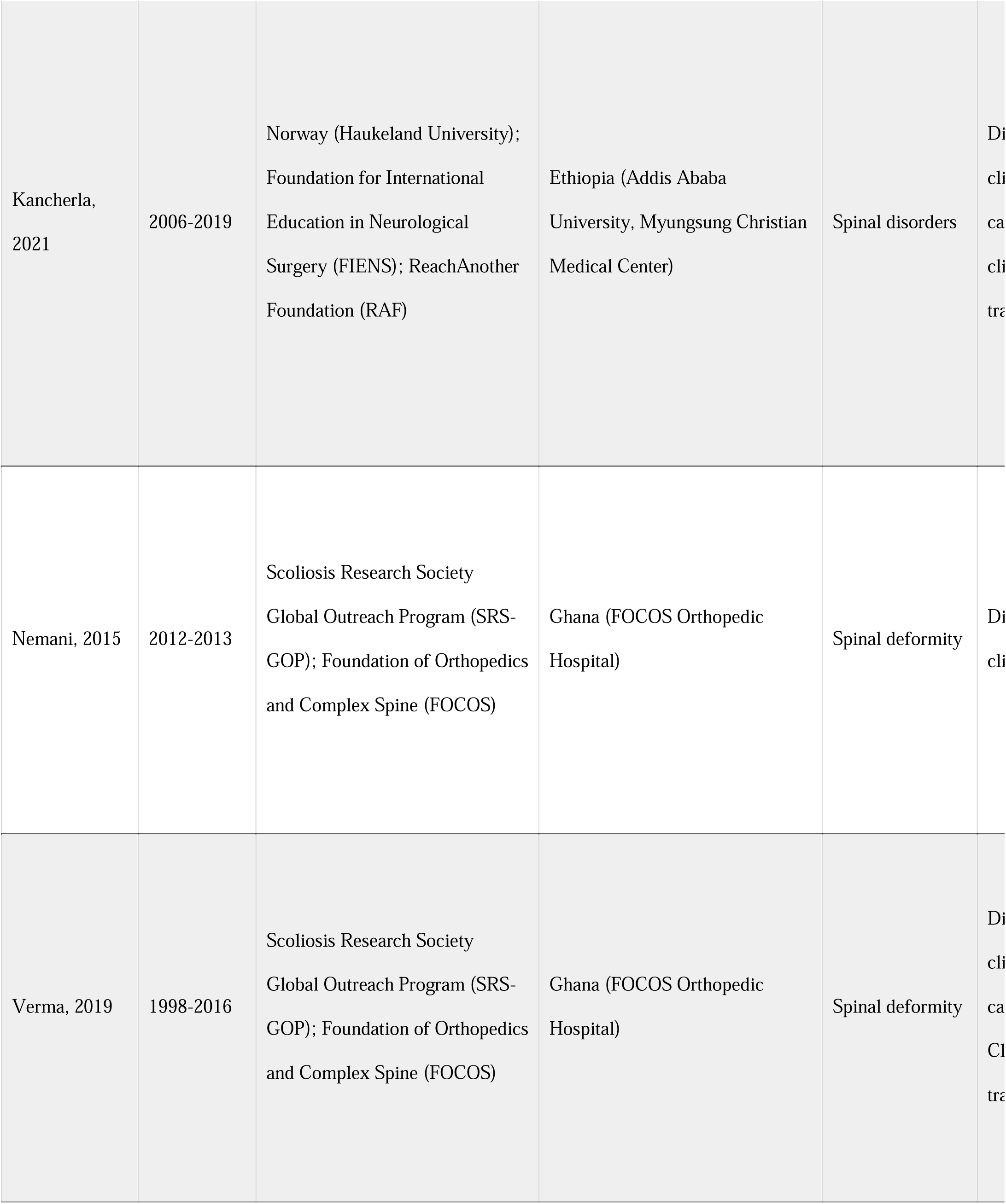
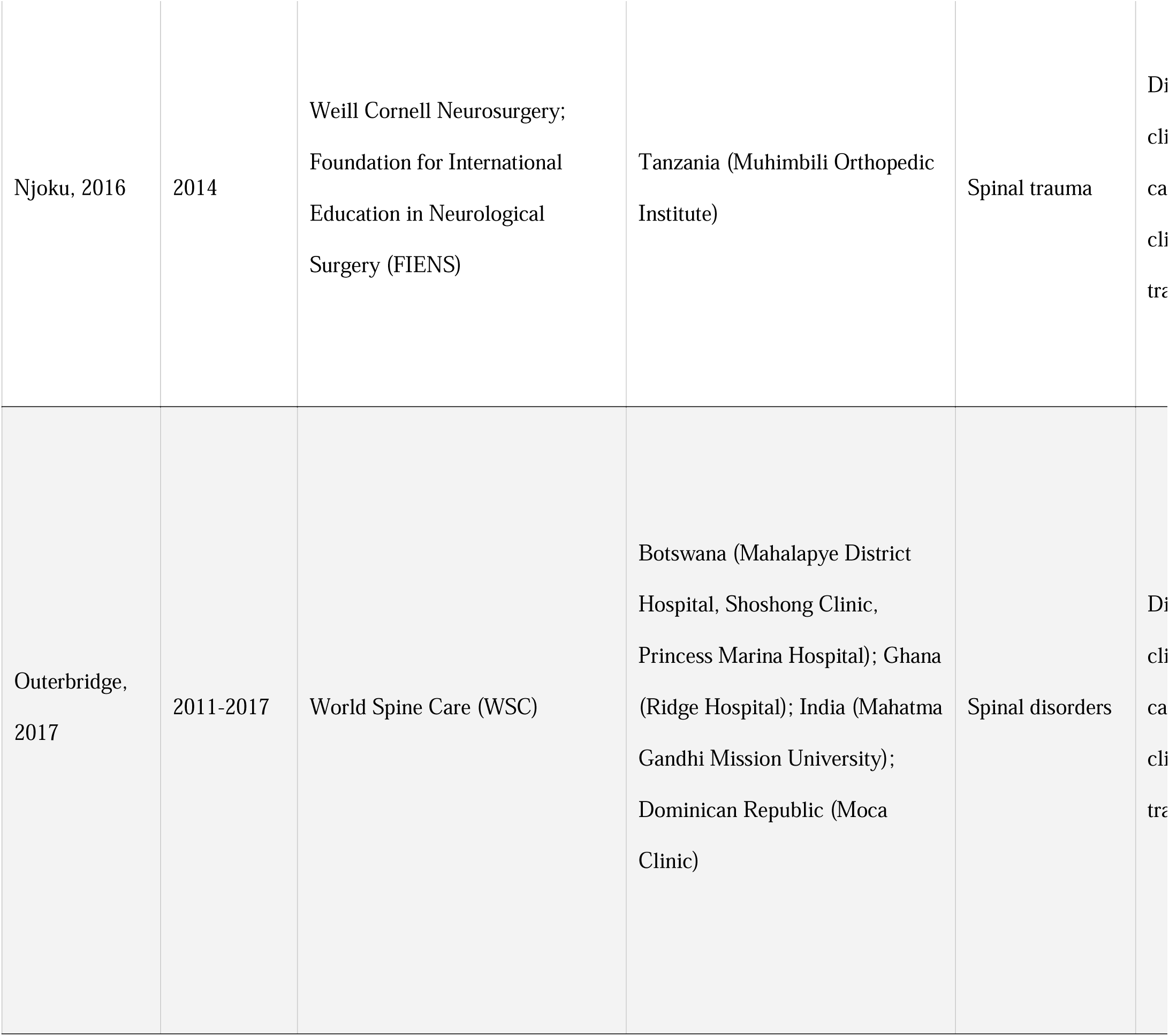
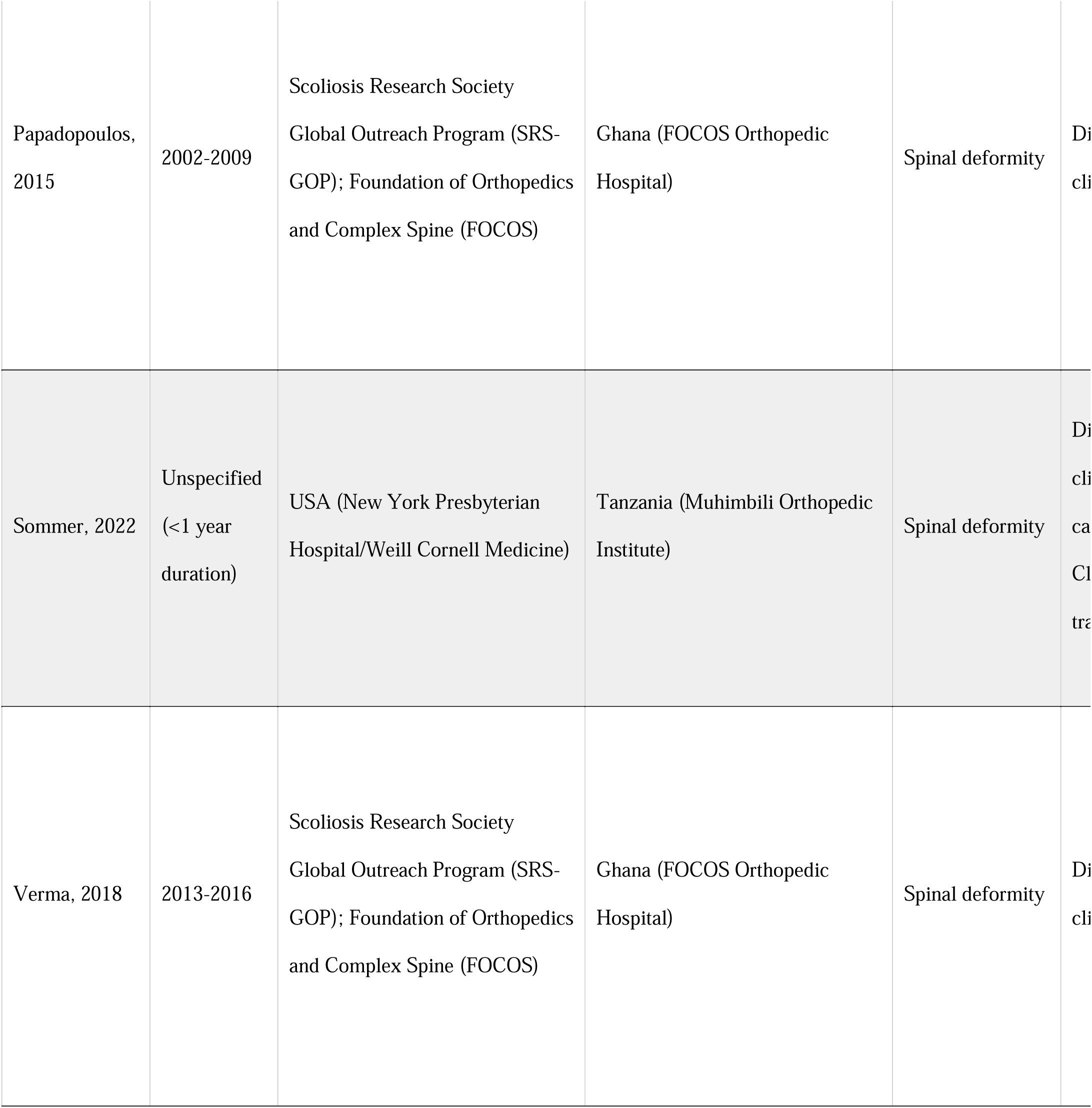
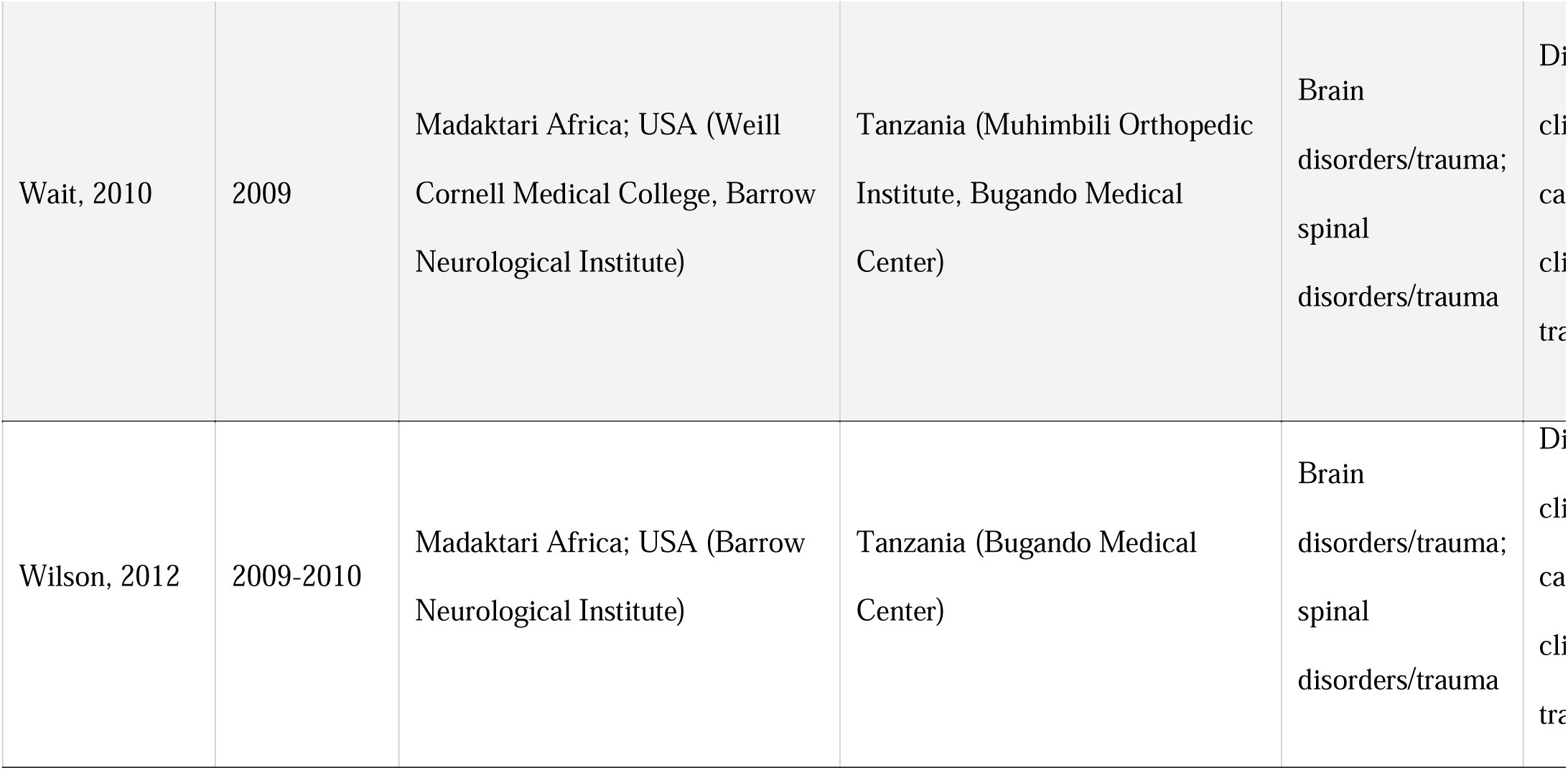
Extracted details from final studies.

| First author,<br>Publication<br>year | Reported<br>date range<br>of study<br>partnership | High resource<br>country/organization/institution | Low resource<br>country/organization/institution | Reported<br>partnership<br>primary focus | Re<br>cli<br>pa<br>ac |
| --- | --- | --- | --- | --- | --- |
| Ahmad, 2023 | Unspecified<br>(<1 year<br>duration) | Weill Cornell Global<br>Neurosurgery Initiative | College of Surgeons of East,<br>Central and Southern Africa<br>(COSECSA) | Spinal deformity | Di<br>cli<br>ca<br>cli<br>tra |
| Boachie-Adjei, 2014 | Unspecified | Scoliosis Research Society<br>Global Outreach Program (SRS-GOP); Foundation of Orthopedics and Complex Spine (FOCOS) | Ghana (FOCOS Orthopedic Hospital) | Spinal deformity | Di<br>cli |
| Boachie-Adjei, 2015 | 1998-2012 | Scoliosis Research Society<br>Global Outreach Program (SRS-GOP); Foundation of Orthopedics and Complex Spine (FOCOS) | Ghana (FOCOS Orthopedic Hospital) | Spinal deformity | Di<br>cli |
| Brady, 2016 | 2009-2016 | World Spine Care (WSC) | Botswana (Mahalapye District Hospital, Shoshong Clinic);<br>Dominican Republic (Moca Clinic) | Spinal disorders | Di<br>cli<br>ca<br>Cl<br>tra |
| Chihambakwe, 2019 | 2011-2017 | World Spine Care (WSC) | Botswana (Mahalapye District Hospital) | Spinal disorders | Di<br>cli |
| Coburger,<br>2014 | 2009-2011 | Madaktari Africa; USA;<br>Germany | Tanzania (Bugando Medical<br>Center) | Brain<br>disorders/trauma;<br>spinal<br>disorders/trauma | Di<br>cli<br>ca<br>Cl<br>tra |
| Fletcher, 2019 | 2008-2016 | Scoliosis Research Society<br>Global Outreach Program (SRS-GOP); Project Perfect World<br>(PPW) | Ecuador (Roberto Gilbert<br>Elizalde Children's Hospital) | Spinal deformity | Di<br>cli<br>ca<br>Cl<br>tra |
| Haldeman,<br>2015 | 2012-2014 | World Spine Care (WSC) | Botswana (Mahalapye District<br>Hospital, Shoshong Clinic);<br>Dominican Republic | Spinal disorders | Di<br>cli<br>ca<br>cli<br>tra |
| Kahamba,<br>2013 | 2011 | USA; Spain; Turkey | Tanzania (Muhimbili Orthopedic<br>Institute); India | Brain<br>disorders/trauma;<br>spinal<br>disorders/trauma | Di<br>cli<br>ca<br>cli<br>tra |
| Kancherla, 2021 | 2006-2019 | Norway (Haukeland University);<br>Foundation for International<br>Education in Neurological<br>Surgery (FIENS); ReachAnother<br>Foundation (RAF) | Ethiopia (Addis Ababa<br>University, Myung Sung Christian<br>Medical Center) | Spinal disorders | Di<br>cli<br>ca<br>cli<br>tra |
| Nemani, 2015 | 2012-2013 | Scoliosis Research Society<br>Global Outreach Program (SRS-GOP); Foundation of Orthopedics<br>and Complex Spine (FOCOS) | Ghana (FOCOS Orthopedic<br>Hospital) | Spinal deformity | Di<br>cli |
| Verma, 2019 | 1998-2016 | Scoliosis Research Society<br>Global Outreach Program (SRS-GOP); Foundation of Orthopedics<br>and Complex Spine (FOCOS) | Ghana (FOCOS Orthopedic<br>Hospital) | Spinal deformity | Di<br>cli<br>ca<br>Cl<br>tra |
| Njoku, 2016 | 2014 | Weill Cornell Neurosurgery;<br>Foundation for International<br>Education in Neurological<br>Surgery (FIENS) | Tanzania (Muhimbili Orthopedic<br>Institute) | Spinal trauma | Di<br>cli<br>ca<br>cli<br>tra |
| Outerbridge,<br>2017 | 2011-2017 | World Spine Care (WSC) | Botswana (Mahalapye District<br>Hospital, Shoshong Clinic,<br>Princess Marina Hospital); Ghana<br>(Ridge Hospital); India (Mahatma<br>Gandhi Mission University);<br>Dominican Republic (Moca<br>Clinic) | Spinal disorders | Di<br>cli<br>ca<br>cli<br>tra |
| Papadopoulos, 2015 | 2002-2009 | Scoliosis Research Society<br>Global Outreach Program (SRS-GOP); Foundation of Orthopedics and Complex Spine (FOCOS) | Ghana (FOCOS Orthopedic Hospital) | Spinal deformity | Di<br>cli |
| Sommer, 2022 | Unspecified<br>($<1$ year duration) | USA (New York Presbyterian Hospital/Weill Cornell Medicine) | Tanzania (Muhimbili Orthopedic Institute) | Spinal deformity | Di<br>cli<br>ca<br>Cl<br>tra |
| Verma, 2018 | 2013-2016 | Scoliosis Research Society<br>Global Outreach Program (SRS-GOP); Foundation of Orthopedics and Complex Spine (FOCOS) | Ghana (FOCOS Orthopedic Hospital) | Spinal deformity | Di<br>cli |
| Wait, 2010 | 2009 | Madaktari Africa; USA (Weill Cornell Medical College, Barrow Neurological Institute) | Tanzania (Muhimbili Orthopedic Institute, Bugando Medical Center) | Brain disorders/trauma; spinal disorders/trauma | Di<br>cli<br>ca<br>cli<br>tra |
| Wilson, 2012 | 2009-2010 | Madaktari Africa; USA (Barrow Neurological Institute) | Tanzania (Bugando Medical Center) | Brain disorders/trauma; spinal disorders/trauma | Di<br>cli<br>ca<br>cli<br>tra |

### Dates and settings of partnerships

Although our literature search encompassed articles published from 2000 until early 2023, all of the studies that fit the final inclusion criteria (19/19, 100%) were published in or after 2010. All reported study partnerships took place between 1998 and 2019. 10/19 (53%) of the articles reported on long-term partnerships with a duration of three years or more.

The included articles revealed four leading global outreach organizations that served as HR partners in their respective collaborations. The SRS-GOP and the Foundation of Orthopedics and Complex Spine (FOCOS), two organizations focused on spinal deformity care and education of local surgeons, were involved as HR partners together in six studies (32%) [12, 14, 16, 18, 20, 23]. SRS-GOP also featured in one other article as a HR partner alongside Project Perfect World (PPW), an organization aiming to improve pediatric orthopedic care in Ecuador [17]. WSC, an organization providing evidence-based spine care to LR communities, was the reported HR partner in four articles (21%) [1, 10, 25, 27]. Three articles (16%) involved Madaktari Africa, an organization dedicated to training healthcare workers in Sub-Saharan Africa, as an HR partner [15, 26, 28]. Other organizations featured as HR partners were the Foundation for International Education in Neurological Surgery (FIENS) and ReachAnotherFoundation (RAF) [19, 21]. Three studies (16%) did not report affiliations with any specific global outreach organizations, instead involving individuals or teams of surgeons from hospitals in various HR countries [13, 22, 24].

Reported LR partners spanned across four global regions: Africa (18/19, 95%) [1, 10, 12–16, 18– 28], the Caribbean (3/19, 16%) [10, 25], South Asia (2/19, 11%) [24, 27], and South America (1/19, 5%) [17]. Interventions were most concentrated in Africa, where Ghana (7/19, 37%) [12, 14, 16, 18, 20, 23, 27] was the most frequently involved LR partner country, while Tanzania (6/19, 32%) [13, 15, 19, 24, 26, 28] and Botswana (4/19, 21%) [1, 10, 25, 27] were also involved in multiple partnerships. All six articles in which SRS-GOP and FOCOS worked together as HR partners centered around care provided at the FOCOS Orthopedic Hospital in Accra, Ghana [12, 14, 16, 18, 20, 23]. WSC partnerships occurred in Botswana, Dominican Republic, Ghana, and India [1, 10, 25, 27]. Partnerships based in Tanzania took place at two different institutions: Bugando Medical Center in Mwanza [15, 26, 28] and Muhimbili Orthopedic Hospital in Dar es Salaam [13, 19, 24, 28].

### Reported partnership focuses, activities, and participants

Nine studies (47%) [12–14, 16–18, 20, 22, 23] focused specifically on spinal deformity care, while five (26%) [1, 10, 21, 25, 27] focused on general spinal disorders, one (5%) [19] focused on spinal trauma, and four (21%) [15, 24, 26, 28] focused on a combination of brain and spinal disorders and trauma.

All of the partnerships included direct clinical care of patients (19/19, 100%). A slight majority of the studies (13/19, 68%) involved clinical training of healthcare providers by HR country physicians. All of the studies (19/19, 100%) involved physicians and physicians-in-training (e.g. residents, medical students) as partnership participants. Eight of the studies (42%) also included non-physician participants, such as nursing staff [16, 24, 28], research assistants [14, 28], chiropractors and physiotherapists [1, 17, 25, 27], and other clinic staff [17].

### Reported needs assessments

A majority of the reviewed articles (14/19, 74%) did not report needs assessments (e.g. literature reviews, focused assessments, interviews with stakeholders) prior to beginning their programs. The five articles that reported needs assessments used various strategies: Coburger et al. [15] and Fletcher and Schwend [17] both reported initial trips to partnership sites intended to assess surgical need, feasibility of complex surgical procedures, and patient population characteristics, while Brady et al. [10] reported a search of existing spine surgery training programs available globally to assess need for surgical trainees. Ahmad et al. [22] utilized needs assessment surveys before, during, and after their training course to plan content and analyze course efficacy, and Haldeman et al. [25] described their assessment of existing facilities prior to officially setting up a clinic.

### Main evaluation focuses and tools

The reviewed articles are split in terms of evaluation focus: twelve studies (63%) [1, 10, 15, 17, 18, 21, 22, 24–28] evaluated some aspect of the partnership itself (e.g. development, effectiveness of training initiative, challenges, local perceptions), six studies (32%) [12, 14, 16, 17, 20, 23] evaluated the clinical outcomes of the care provided through their partnerships, and two studies (11%) [13, 19] evaluated the feasibility of a technological system for surgical assistance.

The articles that evaluated aspects of their partnerships utilized mostly narrative and other qualitative approaches, such as reflections [10, 17, 18, 25–28] and interviews [1]. The five studies that evaluated the efficacy of their partnerships’ surgical training initiatives utilized a mixture of quantitative and qualitative assessment tools, such as surveys, patient outcomes, and narrative reflections [15, 21, 22, 24, 26]. The articles that evaluated clinical outcomes utilized largely quantitative tools such as radiographic measures and clinical indicators [12, 14, 16, 17, 20, 23], and three of these articles also utilized the SRS-22, a validated scoliosis patient-reported outcome questionnaire [16, 17, 20].

## Discussion

To the authors’ knowledge, this is the first review to present and analyze the available literature reporting on spine-based clinical partnerships between HR and LR countries and organizations. Overall, this review identified several HR outreach organizations that are consistently involved in clinical spine partnerships, but there was significant variety in the evaluative focus of published articles and a relative paucity of peer-reviewed articles reporting on partnerships despite the long durations of some interventions.

### Large focus on spinal deformity over other spine care needs in majority of partnerships

In LR countries, traumatic spine injury (TSI) and degenerative spine disease are widely reported as significant needs. Low-and middle-income countries carry a heavier burden of TSI than high-income countries, with an incidence of 13.7 per 100,000 people per year compared to 8.7 per 100,000 people per year [29]. Researchers have published numerous studies on TSI and spinal cord injury (SCI) in LR countries in East and Sub-Saharan Africa and Southeast Asia [31–34]. An analysis of ten years of spine surgery patients at a Nigerian hospital showed that degenerative spine disease was the most common indicator of surgery, accounting for an overwhelming 52.3% of all cases [35].

Given these data, it appears that the reviewed articles show an overrepresentation of spinal deformity care as a focus of HR-LR country partnerships. Only five of the 19 reviewed studies report surgical care of TSI and degenerative spine disease [15, 19, 24, 26, 28]. The reviewed studies show that several organizations are already dominating the spinal deformity field in LR countries, but Madaktari Africa seems to be the only organization regularly publishing studies on TSI or degenerative spine disease. Those considering expanding or initiating partnerships in the future may find it helpful to conduct formal needs assessments at LR sites to consider targeting partnership resources toward conditions most closely matching clinical needs.

### Balance between direct clinical care and capacity building as main reported partnership activities

All reviewed studies reported direct clinical care of patients, and a majority also reported clinical training of local physicians—a significant component of capacity building in a LR country. Several research groups have pinpointed the essential role of capacity building as the guiding goal of ethical global surgery initiatives in order to justify the involvement of outreach volunteers from HR countries [27–29]. Although direct clinical care is a crucial immediate need to address, it may be most effective when it is accompanied by clinical training of local physicians, with the long-term goal of strengthening local health systems and transferring full ownership of sustainable programs to local healthcare providers [25]. Future publications on spine care partnerships should continue to report the specific efforts taken to sustainably strengthen health systems, particularly provider education.

### Varied focuses and tools of partnership evaluation

A majority of the reviewed articles focused on evaluating some aspect of the development, efficacy, or perceptions of the partnership. The rest of the articles focused on either evaluating the clinical outcomes of the care provided by partner surgeons or the technology used during the partnership; in essence, these studies had already accepted the partnership as an established environment for clinical care to be carried out and therefore did not evaluate the nature of the partnership itself.

Although some articles that evaluated the efficacy of their training initiatives utilized surveys for feedback and assessment, there is room for more established frameworks to be used, such as the Kirkpatrick [39], REAIM [40], or CFIR [41] methods, which are already regularly utilized to evaluate training and education interventions. For studies that focus on evaluating training initiatives [15, 21, 22, 24, 26], standardized tools may prove additionally useful in the future. More formal frameworks may be employed to facilitate comparison with other initiatives or specialties and may guide future steps to ensure evidence-based improvement.

### Promising needs assessments reported

Most of the reviewed articles did not report any needs assessments at their planned sites prior to the start of their programs. However, the five studies that reported needs assessments demonstrate feasible steps for future program partners to build on in order to plan ethical and effective partnerships: systematic literature searches, surveys, discussions with local hospital staff, and preliminary trips to the partnership site. This crucial step allows partners to mutually define a clear goal and scope of their program based on a current comprehensive evaluation of clinical and structural needs.

Various groups in other surgical specialties have published their work on developing needs assessments to build a foundation for their outreach programs, providing other models that future spine-based partnerships might also consider [42, 43]. Future spine clinical care partnerships should conduct and report comprehensive qualitative and quantitative needs assessments with involved stakeholders through interviews, focus groups, observational studies, focused surveys, and other evaluative methods to provide a strong foundation for the development of an evidence-based partnership.

### Opportunity for increased reporting on spine-based clinical partnerships

Despite an extensive literature search, only nineteen studies qualified for final review. Other reviews surveying the current state of global clinical partnerships in anesthesiology [37] and trauma [38] have found at least two times as many qualifying articles, signaling that spine-based clinical partnerships are not as well-reported in peer-reviewed literature. There is good reason to believe that this results from under-reporting of existing partnerships: for instance, an internet search for spine global partnerships returns various websites of organizations already included in this review, but other organizations with accessible online evidence of functioning partnerships also appear (e.g., Butterfly Foundation Spine, Global Spine Outreach). It is likely that these organizations are having a substantial impact on clinical care, and it would be helpful to have more accessible information in the literature about their impact and approaches.

Although the nineteen studies reviewed in this paper revealed that some of these organizations already work together, such as SRS and FOCOS, it is possible that the concentrated areas in which these programs tend to operate (e.g. Ghana, Tanzania) may have already generated other unreported collaborations between organizations. Because spine care uniquely unites two surgical specialties— orthopedic surgery and neurosurgery—as well as non-surgical specialists like physiatrists and chiropractors, there is immense potential for interdisciplinary collaboration. Increasing publication on partnership development, implementation, and outcomes could aid in awareness that in turn stimulates further collaboration.

To ensure optimal knowledge of the efforts being taken by HR spine care outreach groups in LR settings, it may be useful for authors to report the nature of their collaborations in all published articles, even if the partnership itself is not the focal point of their study. In this way, articles arising from even informal collaborations may still be used as a foundation for improved, effective partnerships in the future.

### Limitations

This review was only able to analyze studies on spine-based clinical partnerships published in three online research databases. Many rejected articles included authors representing both HR institutions and LR institutions but did not report any partnership details in their text [31, 32, 34]. The existence of these articles illuminates the prevalence of peer-reviewed articles produced by HR-LR partnerships that either are not formalized or focus purely on non-clinical research collaborations, leaving room for further analysis of the larger scope of informal global spine care partnerships operating without the involvement of large outreach organizations.

Additionally, since the search strategy was conducted in English, articles in different languages may have been excluded. Therefore, more articles on these partnerships may exist that were not identified and the reported findings should be interpreted with these limitations in mind.

## Conclusion

To the authors’ knowledge, this scoping review is the first study to search for and analyze the current literature available on spine-based clinical partnerships between HR and LR countries. Overall, this review revealed the relative scarcity of published studies on global spine clinical care partnerships despite the clear presence and continued work of many HR global outreach organizations in LR countries. The current studies varied in their evaluation focuses, but the articles that evaluated aspects of their partnerships showed promising needs assessments and capacity building efforts. We recommend an increase in the quantity of formal evaluations and peer-reviewed studies on spine-based clinical partnerships to inform the successful, equitable, and accountable implementation of future partnerships and to promote quality spine care worldwide.

## Data Availability

All relevant data are within the manuscript and its Supporting Information files.

## Supporting information

S1 Appendix. Scoping review search strategy.

S2 Table. PRISMA-ScR checklist.

