## Supplement 1 for "Clinical spine care partnerships between low- and high-resource countries: A scoping review"

**PubMed**

(“Global Health”[Mesh] OR “Global health”[tw] OR humanitarian[tw] OR “Medical Missions”[mesh] OR “medical mission”[tw] OR “medical missions”[tw] OR volunteers[mesh] OR volunteers[tw] OR volunteer[tw] OR outreach[tw] OR health equity[mesh] OR international cooperation[mesh] OR organizations, nonprofit[mesh] OR Global outreach[tw] OR world spine care[tiab] OR international program[tiab] OR ((Afghanistan[Affiliation] OR Albania[Affiliation] OR Algeria[Affiliation] OR Angola[Affiliation] OR Argentina[Affiliation] OR Armenia[Affiliation] OR Azerbaijan[Affiliation] OR Bangladesh[Affiliation] OR Belarus[Affiliation] OR Belize[Affiliation] OR Benin[Affiliation] OR Bhutan[Affiliation] OR Bolivia[Affiliation] OR Bosnia and Herzegovina[Affiliation] OR Botswana[Affiliation] OR Brazil[Affiliation] OR Burkina Faso[Affiliation] OR Burundi[Affiliation] OR Cabo Verde[Affiliation] OR Cambodia[Affiliation] OR Cameroon[Affiliation] OR Central African Republic[Affiliation] OR Chad[Affiliation] OR People's Republic of China [Affiliation] OR Colombia[Affiliation] OR Comoros[Affiliation] OR Democratic Republic of Congo[Affiliation] OR Congo[Affiliation] OR Costa Rica[Affiliation] OR Côte d'Ivoire[Affiliation] OR Cuba[Affiliation] OR Djibouti[Affiliation] OR Dominica[Affiliation] OR Dominican Republic[Affiliation] OR Ecuador[Affiliation] OR Egypt[Affiliation] OR El Salvador[Affiliation] OR Equatorial Guinea[Affiliation] OR Eritrea[Affiliation] OR Eswatini[Affiliation] OR Ethiopia[Affiliation] OR Fiji[Affiliation] OR Gabon[Affiliation] OR Gambia[Affiliation] OR Georgia[Affiliation] OR Ghana[Affiliation] OR Grenada[Affiliation] OR Guatemala[Affiliation] OR Guinea[Affiliation] OR Guinea-Bissau[Affiliation] OR Guyana[Affiliation] OR Haiti[Affiliation] OR Honduras[Affiliation] OR India[Affiliation] OR Indonesia[Affiliation] OR Iran[Affiliation] OR Iraq[Affiliation] OR Jamaica[Affiliation] OR Jordan[Affiliation] OR Kazakhstan[Affiliation] OR Kenya[Affiliation] OR Kiribati[Affiliation] OR Democratic People's Republic of Korea[Affiliation] OR Kosovo[Affiliation] OR Kyrgyzstan[Affiliation] OR Lao People's Democratic Republic[Affiliation] OR Lebanon[Affiliation] OR Lesotho[Affiliation] OR Liberia[Affiliation] OR Libya[Affiliation] OR North Macedonia[Affiliation] OR Madagascar[Affiliation] OR Malawi[Affiliation] OR Malaysia[Affiliation] OR Maldives[Affiliation] OR Mali[Affiliation] OR Marshall Islands[Affiliation] OR Mauritania[Affiliation] OR Mauritius[Affiliation] OR Mexico[Affiliation] OR Micronesia[Affiliation] OR Moldova[Affiliation] OR Mongolia[Affiliation] OR Montenegro[Affiliation] OR Montserrat[Affiliation] OR Morocco[Affiliation] OR Mozambique[Affiliation] OR Myanmar[Affiliation] OR Namibia[Affiliation] OR Nauru[Affiliation] OR Nepal[Affiliation] OR Nicaragua[Affiliation] OR Niger[Affiliation] OR Nigeria[Affiliation] OR Niue[Affiliation] OR Pakistan[Affiliation] OR Panama[Affiliation] OR Papua New Guinea[Affiliation] OR Paraguay[Affiliation] OR Peru[Affiliation] OR Philippines[Affiliation] OR Rwanda[Affiliation] OR Saint Helena[Affiliation] OR Samoa[Affiliation] OR São Tomé and Príncipe[Affiliation] OR Senegal[Affiliation] OR Serbia[Affiliation] OR Sierra Leone[Affiliation] OR Solomon Islands[Affiliation] OR Somalia[Affiliation] OR South Africa[Affiliation] OR South Sudan[Affiliation] OR Sri Lanka[Affiliation] OR Saint Lucia[Affiliation] OR Saint Vincent and the Grenadines[Affiliation] OR Sudan[Affiliation] OR Suriname[Affiliation] OR Syrian Arab Republic[Affiliation] OR Tajikistan[Affiliation] OR Tanzania[Affiliation] OR Thailand[Affiliation] OR Timor-Leste[Affiliation] OR Togo[Affiliation] OR Tokelau[Affiliation] OR Tonga[Affiliation] OR Tunisia[Affiliation] OR Turkey[Affiliation] OR Turkmenistan[Affiliation] OR Tuvalu[Affiliation] OR Uganda[Affiliation] OR Ukraine[Affiliation] OR Uzbekistan[Affiliation] OR Vanuatu[Affiliation] OR Venezuela[Affiliation] OR Vietnam[Affiliation] OR Wallis Futuna[Affiliation] OR West Bank Gaza Strip[Affiliation] OR Yemen[Affiliation] OR Zambia[Affiliation] OR Zimbabwe[Affiliation]) AND (Alabama[Affiliation] OR Alaska[Affiliation] OR Arizona[Affiliation] OR Arkansas[Affiliation] OR California[Affiliation] OR Colorado[Affiliation] OR Connecticut[Affiliation] OR Delaware[Affiliation] OR Florida[Affiliation] OR Georgia[Affiliation] OR Hawaii[Affiliation] OR Idaho[Affiliation] OR Illinois[Affiliation] OR Indiana[Affiliation] OR Iowa[Affiliation] OR Kansas[Affiliation] OR Kentucky[Affiliation] OR Louisiana[Affiliation] OR Maine[Affiliation] OR Maryland[Affiliation] OR Massachusetts[Affiliation] OR Michigan[Affiliation] OR Minnesota[Affiliation] OR Mississippi[Affiliation] OR Missouri[Affiliation] OR Montana[Affiliation] OR Nebraska[Affiliation] OR Nevada[Affiliation] OR New Hampshire[Affiliation] OR New Jersey[Affiliation] OR New Mexico[Affiliation] OR New York[Affiliation] OR North Carolina[Affiliation] OR North Dakota[Affiliation] OR Ohio[Affiliation] OR Oklahoma[Affiliation] OR Oregon[Affiliation] OR Pennsylvania[Affiliation] OR Rhode Island[Affiliation] OR South Carolina[Affiliation] OR South Dakota[Affiliation] OR Tennessee[Affiliation] OR Texas[Affiliation] OR Utah[Affiliation] OR Vermont[Affiliation] OR Virginia[Affiliation] OR Washington[Affiliation] OR West Virginia[Affiliation] OR Wisconsin[Affiliation] OR Wyoming[Affiliation] OR United States[Affiliation] OR USA[Affiliation] OR Canada[Affiliation] OR Japan[Affiliation] OR United Kingdom[Affiliation] OR Germany[Affiliation] OR France[Affiliation] OR Australia[Affiliation] OR Italy[Affiliation] OR South Korea[Affiliation] OR Spain[Affiliation] OR Sweden[Affiliation] OR Switzerland[Affiliation] OR Norway[Affiliation] OR Netherlands[Affiliation] OR Austria[Affiliation] OR Belgium[Affiliation] OR Denmark[Affiliation] OR Finland[Affiliation] OR Ireland[Affiliation] OR Iceland[Affiliation] OR Croatia[Affiliation] OR Czech Republic[Affiliation] OR New Zealand[Affiliation] OR Hungary[Affiliation] OR Poland[Affiliation] OR Singapore[Affiliation])))

AND

(education, medical[Mesh] OR Education[tw] OR Research[Mesh] OR research[tw] OR training[tw] OR evaluation[tw] OR analysis[tw] OR "Needs Assessment"[Mesh] OR needs assessment[tw] OR "Outcome Assessment, Health Care"[Mesh] OR observe[tw] OR survey[tw] OR interview[tw] OR audit[tw] OR checklist[tw] OR measurement[tw] OR results[tw] OR improvement[tw] OR impact[tw] OR benefit[tw] OR effect[tw] OR strengthen[tw] OR build[tw] OR improve[tw] OR clinical[tw] OR clinic[tw] OR clinics[tw] OR "Patient-Centered Care"[Mesh] OR patient care[tw])

AND

(“resource limited”[tw] OR “limited resource”[tw] OR “resource poor”[tw] OR underserved region[tw] OR “low-to-middle-income country”[tw] OR “low-to-middle-income countries”[tw] OR “low-and-middle-income country”[tw] OR “low-and-middle-income countries”[tw] OR “low-income”[tw] OR “Low Resource”[tw] OR “third-world”[tw] OR “Developing Countries”[mesh] OR “Developing country”[tw] OR “Developing countries”[tw] OR “Developing Nation”[tw] OR “Developing Nations”[tw] OR “developing population”[tw] OR “developing populations”[tw] OR Africa South of the Sahara[Mesh] OR sub-saharan[tw] OR sub-sahara[tw] OR south of the Sahara[tw] OR West Africa[tw] OR Western Africa[tw] OR Benin[tw] OR Burkina Faso[tw] OR Cabo Verde[tw] OR Cape Verde[tw] OR Cote d'Ivoire[tw] OR Ivory Coast[tw] OR Gambia[tw] OR Ghana[tw] OR Guinea[tw] OR Guinea-Bissau[tw] OR Liberia[tw] OR Mali[tw] OR Mauritania[tw] OR Niger[tw] OR Nigeria[tw] OR Senegal[tw] OR Sierra Leone[tw] OR Togo[tw] OR Central Africa[tw] OR Cameroon[tw] OR Central African Republic[tw] OR Chad[tw] OR Congo[tw] OR Democratic Republic of the Congo[tw] OR Equatorial Guinea[tw] OR Gabon[tw] OR “Sao Tome and Principe”[tw] OR Eastern Africa[tw] OR Burundi[tw] OR Djibouti[tw] OR Eritrea[tw] OR Ethiopia[tw] OR Kenya[tw] OR Rwanda[tw] OR Somalia[tw] OR South Sudan[tw] OR Sudan[tw] OR Tanzania[tw] OR Uganda[tw] OR Southern Africa[tw] OR Angola[tw] OR Botswana[tw] OR Eswatini[tw] OR Lesotho[tw] OR Malawi[tw] OR Mozambique[tw] OR Namibia[tw] OR South Africa[tw] OR Zambia[tw] OR Zimbabwe[tw] OR Asia, Southeastern[mesh] OR Southeast Asia[tw] OR Borneo[tw] OR Brunei[tw] OR Cambodia[tw] OR Indochina[tw] OR Indonesia[tw] OR Laos[tw] OR Malaysia[tw] OR Mekong Valley[tw] OR Myanmar[tw] OR Philippines[tw] OR Singapore[tw] OR Thailand[tw] OR Timor-Leste[tw] OR Vietnam[tw] OR Solomon Islands[tw])

AND

("Spine"[Mesh] OR Spine[tw] OR spines[tw] OR spinal[tw] OR Vertebral[tw] OR Vertebra[tw] OR Vertebrae[tw] OR thoracic[tw] OR lumbar[tw] OR sacrum[tw] OR intervertebral[tw])

NOT (animals[mesh] NOT humans[mesh]) AND (2000/1/1:3000/12/12[pdat])

**Embase**

("Global Health"/exp OR "Global health":ti,ab,de,tn,kw OR “humanitarian”:ti,ab,de,tn,kw OR 'international cooperation'/exp OR "medical mission":ti,ab,de,tn,kw OR "medical missions":ti,ab,de,tn,kw OR volunteer/exp OR “volunteers”:ti,ab,de,tn,kw OR “volunteer”:ti,ab,de,tn,kw OR “outreach”:ti,ab,de,tn,kw OR "health equity"/exp OR "international cooperation"/exp OR 'non profit organization'/exp OR 'Global outreach':ti,ab,de,tn,kw OR 'world spine care':ti,ab OR 'international program':ti,ab OR ((Afghanistan:ca OR Albania:ca OR Algeria:ca OR Angola:ca OR Argentina:ca OR Armenia:ca OR Azerbaijan:ca OR Bangladesh:ca OR Belarus:ca OR Belize:ca OR Benin:ca OR Bhutan:ca OR Bolivia:ca OR ‘Bosnia and Herzegovina’:ca OR Botswana:ca OR Brazil:ca OR ‘Burkina Faso’:ca OR Burundi:ca OR ‘Cabo Verde’:ca OR Cambodia:ca OR Cameroon:ca OR ‘Central African Republic’:ca OR Chad:ca OR ‘Peoples Republic of China’:ca OR Colombia:ca OR Comoros:ca OR ‘Democratic Republic of Congo’:ca OR Congo:ca OR ‘Costa Rica’:ca OR ‘Cote d Ivoire’:ca OR Cuba:ca OR Djibouti:ca OR Dominica:ca OR ‘Dominican Republic’:ca OR Ecuador:ca OR Egypt:ca OR ‘El Salvador’:ca OR ‘Equatorial Guinea’:ca OR Eritrea:ca OR Eswatini:ca OR Ethiopia:ca OR Fiji:ca OR Gabon:ca OR Gambia:ca OR Georgia:ca OR Ghana:ca OR Grenada:ca OR Guatemala:ca OR Guinea:ca OR Guinea-Bissau:ca OR Guyana:ca OR Haiti:ca OR Honduras:ca OR India:ca OR Indonesia:ca OR Iran:ca OR Iraq:ca OR Jamaica:ca OR Jordan:ca OR Kazakhstan:ca OR Kenya:ca OR Kiribati:ca OR ‘Democratic Peoples Republic of Korea’:ca OR Kosovo:ca OR Kyrgyzstan:ca OR ‘Lao Peoples Democratic Republic’:ca OR Lebanon:ca OR Lesotho:ca OR Liberia:ca OR Libya:ca OR ‘North Macedonia’:ca OR Madagascar:ca OR Malawi:ca OR Malaysia:ca OR Maldives:ca OR Mali:ca OR ‘Marshall Islands’:ca OR Mauritania:ca OR Mauritius:ca OR Mexico:ca OR Micronesia:ca OR Moldova:ca OR Mongolia:ca OR Montenegro:ca OR Montserrat:ca OR Morocco:ca OR Mozambique:ca OR Myanmar:ca OR Namibia:ca OR Nauru:ca OR Nepal:ca OR Nicaragua:ca OR Niger:ca OR Nigeria:ca OR Niue:ca OR Pakistan:ca OR Panama:ca OR ‘Papua New Guinea’:ca OR Paraguay:ca OR Peru:ca OR Philippines:ca OR Rwanda:ca OR ‘Saint Helena’:ca OR Samoa:ca OR ‘São Tomé and Príncipe’:ca OR Senegal:ca OR Serbia:ca OR ‘Sierra Leone’:ca OR ‘Solomon Islands’:ca OR Somalia:ca OR ‘South Africa’:ca OR ‘South Sudan’:ca OR ‘Sri Lanka’:ca OR ‘Saint Lucia’:ca OR ‘Saint Vincent and the Grenadines’:ca OR Sudan:ca OR Suriname:ca OR ‘Syrian Arab Republic’:ca OR Tajikistan:ca OR Tanzania:ca OR Thailand:ca OR Timor-Leste:ca OR Togo:ca OR Tokelau:ca OR Tonga:ca OR Tunisia:ca OR Turkey:ca OR Turkmenistan:ca OR Tuvalu:ca OR Uganda:ca OR Ukraine:ca OR Uzbekistan:ca OR Vanuatu:ca OR Venezuela:ca OR Vietnam:ca OR ‘Wallis Futuna’:ca OR ‘West Bank Gaza Strip’:ca OR Yemen:ca OR Zambia:ca OR Zimbabwe:ca) AND (‘United States’:ca OR USA:ca OR Canada:ca OR Japan:ca OR ‘United Kingdom’:ca OR Germany:ca OR France:ca OR Australia:ca OR Italy:ca OR ‘South Korea’:ca OR Spain:ca OR Sweden:ca OR Switzerland:ca OR Norway:ca OR Netherlands:ca OR Austria:ca OR Belgium:ca OR Denmark:ca OR Finland:ca OR Ireland:ca OR Iceland:ca OR Croatia:ca OR ‘Czech Republic’:ca OR ‘New Zealand’:ca OR Hungary:ca OR Poland:ca OR Singapore:ca)))

AND

('medical education'/exp OR 'education':ti,ab,de,tn,kw OR 'research'/exp OR 'research':ti,ab,de,tn,kw OR 'training':ti,ab,de,tn,kw OR 'evaluation':ti,ab,de,tn,kw OR 'analysis':ti,ab,de,tn,kw OR 'needs assessment'/exp OR 'needs assessment':ti,ab,de,tn,kw OR 'outcome assessment'/exp OR 'observe':ti,ab,de,tn,kw OR 'survey':ti,ab,de,tn,kw OR 'interview':ti,ab,de,tn,kw OR 'audit':ti,ab,de,tn,kw OR 'checklist':ti,ab,de,tn,kw OR 'measurement':ti,ab,de,tn,kw OR 'results':ti,ab,de,tn,kw OR 'improvement':ti,ab,de,tn,kw OR 'impact':ti,ab,de,tn,kw OR 'benefit':ti,ab,de,tn,kw OR 'effect':ti,ab,de,tn,kw OR 'strengthen':ti,ab,de,tn,kw OR 'build':ti,ab,de,tn,kw OR 'improve':ti,ab,de,tn,kw OR “clinical”:ti,ab,de,tn,kw OR “clinic”:ti,ab,de,tn,kw OR “clinics”:ti,ab,de,tn,kw OR 'patient care'/exp)

AND

("resource limited”:ti,ab,de,tn,kw OR "limited resource”:ti,ab,de,tn,kw OR "resource poor”:ti,ab,de,tn,kw OR “underserved region”:ti,ab,de,tn,kw OR "low-to-middle-income country”:ti,ab,de,tn,kw OR "low-to-middle-income countries”:ti,ab,de,tn,kw OR "low-and-middle-income country”:ti,ab,de,tn,kw OR "low-and-middle-income countries”:ti,ab,de,tn,kw OR “low-income”:ti,ab,de,tn,kw OR "Low Resource”:ti,ab,de,tn,kw OR “third-world”:ti,ab,de,tn,kw OR "Developing County"/exp OR "Developing country”:ti,ab,de,tn,kw OR "Developing countries”:ti,ab,de,tn,kw OR "Developing Nation”:ti,ab,de,tn,kw OR "Developing Nations”:ti,ab,de,tn,kw OR "developing population”:ti,ab,de,tn,kw OR "developing populations”:ti,ab,de,tn,kw OR "Africa South of the Sahara"/exp OR “sub-saharan”:ti,ab,de,tn,kw OR “sub-sahara”:ti,ab,de,tn,kw OR "south of the Sahara”:ti,ab,de,tn,kw OR "West Africa”:ti,ab,de,tn,kw OR "Western Africa”:ti,ab,de,tn,kw OR “Benin”:ti,ab,de,tn,kw OR "Burkina Faso”:ti,ab,de,tn,kw OR "Cabo Verde”:ti,ab,de,tn,kw OR "Cape Verde”:ti,ab,de,tn,kw OR "Cote d Ivoire”:ti,ab,de,tn,kw OR "Ivory Coast”:ti,ab,de,tn,kw OR “Gambia”:ti,ab,de,tn,kw OR “Ghana”:ti,ab,de,tn,kw OR “Guinea”:ti,ab,de,tn,kw OR “Guinea-Bissau”:ti,ab,de,tn,kw OR “Liberia”:ti,ab,de,tn,kw OR “Mali”:ti,ab,de,tn,kw OR “Mauritania”:ti,ab,de,tn,kw OR “Niger”:ti,ab,de,tn,kw OR “Nigeria”:ti,ab,de,tn,kw OR “Senegal”:ti,ab,de,tn,kw OR "Sierra Leone”:ti,ab,de,tn,kw OR “Togo”:ti,ab,de,tn,kw OR "Central Africa”:ti,ab,de,tn,kw OR “Cameroon”:ti,ab,de,tn,kw OR "Central African Republic”:ti,ab,de,tn,kw OR “Chad”:ti,ab,de,tn,kw OR “Congo”:ti,ab,de,tn,kw OR "Democratic Republic of the Congo”:ti,ab,de,tn,kw OR "Equatorial Guinea”:ti,ab,de,tn,kw OR “Gabon”:ti,ab,de,tn,kw OR "Sao Tome and Principe”:ti,ab,de,tn,kw OR "Eastern Africa”:ti,ab,de,tn,kw OR “Burundi”:ti,ab,de,tn,kw OR “Djibouti”:ti,ab,de,tn,kw OR “Eritrea”:ti,ab,de,tn,kw OR “Ethiopia”:ti,ab,de,tn,kw OR “Kenya”:ti,ab,de,tn,kw OR “Rwanda”:ti,ab,de,tn,kw OR “Somalia”:ti,ab,de,tn,kw OR "South Sudan”:ti,ab,de,tn,kw OR “Sudan”:ti,ab,de,tn,kw OR “Tanzania”:ti,ab,de,tn,kw OR “Uganda”:ti,ab,de,tn,kw OR "Southern Africa”:ti,ab,de,tn,kw OR “Angola”:ti,ab,de,tn,kw OR “Botswana”:ti,ab,de,tn,kw OR “Eswatini”:ti,ab,de,tn,kw OR “Lesotho”:ti,ab,de,tn,kw OR “Malawi”:ti,ab,de,tn,kw OR “Mozambique”:ti,ab,de,tn,kw OR “Namibia”:ti,ab,de,tn,kw OR "South Africa”:ti,ab,de,tn,kw OR “Zambia”:ti,ab,de,tn,kw OR “Zimbabwe”:ti,ab,de,tn,kw OR 'Southeast Asia'/exp OR "Southeast Asia”:ti,ab,de,tn,kw OR “Borneo”:ti,ab,de,tn,kw OR “Brunei”:ti,ab,de,tn,kw OR “Cambodia”:ti,ab,de,tn,kw OR “Indochina”:ti,ab,de,tn,kw OR “Indonesia”:ti,ab,de,tn,kw OR “Laos”:ti,ab,de,tn,kw OR “Malaysia”:ti,ab,de,tn,kw OR "Mekong Valley”:ti,ab,de,tn,kw OR “Myanmar”:ti,ab,de,tn,kw OR “Philippines”:ti,ab,de,tn,kw OR “Singapore”:ti,ab,de,tn,kw OR “Thailand”:ti,ab,de,tn,kw OR “Timor-Leste”:ti,ab,de,tn,kw OR “Vietnam”:ti,ab,de,tn,kw OR "Solomon Islands”:ti,ab,de,tn,kw)

AND

(Spine/exp OR Spine:ti,ab,de,tn,kw OR spines:ti,ab,de,tn,kw OR spinal:ti,ab,de,tn,kw OR Vertebral:ti,ab,de,tn,kw OR Vertebra:ti,ab,de,tn,kw OR Vertebrae:ti,ab,de,tn,kw OR thoracic:ti,ab,de,tn,kw OR lumbar:ti,ab,de,tn,kw OR sacrum:ti,ab,de,tn,kw OR intervertebral:ti,ab,de,tn,kw)

AND [2000-2023]/py AND [humans]/lim

**Cochrane**

([mh "Global Health"] OR "Global health":ti,ab,kw OR humanitarian:ti,ab,kw OR [mh "Medical Missions"] OR "medical mission":ti,ab,kw OR "medical missions":ti,ab,kw OR [mh volunteers] OR volunteers:ti,ab,kw OR volunteer:ti,ab,kw OR outreach:ti,ab,kw OR [mh "health equity"] OR [mh "international cooperation"] OR [mh "organizations, nonprofit"] OR "Global outreach":ti,ab,kw OR "world spine care":ti,ab OR "international program":ti,ab)

AND

([mh "education, medical"] OR Education:ti,ab,kw OR [mh Research] OR research:ti,ab,kw OR training:ti,ab,kw OR evaluation:ti,ab,kw OR analysis:ti,ab,kw OR [mh "Needs Assessment"] OR "needs assessment":ti,ab,kw OR [mh "Outcome Assessment, Health Care"] OR observe:ti,ab,kw OR survey:ti,ab,kw OR interview:ti,ab,kw OR audit:ti,ab,kw OR checklist:ti,ab,kw OR measurement:ti,ab,kw OR results:ti,ab,kw OR improvement:ti,ab,kw OR impact:ti,ab,kw OR benefit:ti,ab,kw OR effect:ti,ab,kw OR strengthen:ti,ab,kw OR build:ti,ab,kw OR improve:ti,ab,kw OR clinical:ti,ab,kw OR clinic:ti,ab,kw OR clinics:ti,ab,kw OR [mh "Patient-Centered Care"] OR "patient care":ti,ab,kw)

AND

"resource limited":ti,ab,kw OR "limited resource":ti,ab,kw OR "resource poor":ti,ab,kw OR "underserved region":ti,ab,kw OR "low-to-middle-income country":ti,ab,kw OR "low-to-middle-income countries":ti,ab,kw OR "low-and-middle-income country":ti,ab,kw OR "low-and-middle-income countries":ti,ab,kw OR low-income:ti,ab,kw OR "Low Resource":ti,ab,kw OR third-world:ti,ab,kw OR [mh "Developing Countries"] OR "Developing country":ti,ab,kw OR "Developing countries":ti,ab,kw OR "Developing Nation":ti,ab,kw OR "Developing Nations":ti,ab,kw OR "developing population":ti,ab,kw OR "developing populations":ti,ab,kw OR [mh "Africa South of the Sahara"] OR sub-saharan:ti,ab,kw OR sub-sahara:ti,ab,kw OR "south of the Sahara":ti,ab,kw OR "West Africa":ti,ab,kw OR "Western Africa":ti,ab,kw OR Benin:ti,ab,kw OR "Burkina Faso":ti,ab,kw OR "Cabo Verde":ti,ab,kw OR "Cape Verde":ti,ab,kw OR "Cote d'Ivoire":ti,ab,kw OR "Ivory Coast":ti,ab,kw OR Gambia:ti,ab,kw OR Ghana:ti,ab,kw OR Guinea:ti,ab,kw OR Guinea-Bissau:ti,ab,kw OR Liberia:ti,ab,kw OR Mali:ti,ab,kw OR Mauritania:ti,ab,kw OR Niger:ti,ab,kw OR Nigeria:ti,ab,kw OR Senegal:ti,ab,kw OR "Sierra Leone":ti,ab,kw OR Togo:ti,ab,kw OR "Central Africa":ti,ab,kw OR Cameroon:ti,ab,kw OR "Central African Republic":ti,ab,kw OR Chad:ti,ab,kw OR Congo:ti,ab,kw OR "Democratic Republic of the Congo":ti,ab,kw OR "Equatorial Guinea":ti,ab,kw OR Gabon:ti,ab,kw OR "Sao Tome and Principe":ti,ab,kw OR "Eastern Africa":ti,ab,kw OR Burundi:ti,ab,kw OR Djibouti:ti,ab,kw OR Eritrea:ti,ab,kw OR Ethiopia:ti,ab,kw OR Kenya:ti,ab,kw OR Rwanda:ti,ab,kw OR Somalia:ti,ab,kw OR "South Sudan":ti,ab,kw OR Sudan:ti,ab,kw OR Tanzania:ti,ab,kw OR Uganda:ti,ab,kw OR "Southern Africa":ti,ab,kw OR Angola:ti,ab,kw OR Botswana:ti,ab,kw OR Eswatini:ti,ab,kw OR Lesotho:ti,ab,kw OR Malawi:ti,ab,kw OR Mozambique:ti,ab,kw OR Namibia:ti,ab,kw OR "South Africa":ti,ab,kw OR Zambia:ti,ab,kw OR Zimbabwe:ti,ab,kw OR [mh "Asia, Southeastern"] OR "Southeast Asia":ti,ab,kw OR Borneo:ti,ab,kw OR Brunei:ti,ab,kw OR Cambodia:ti,ab,kw OR Indochina:ti,ab,kw OR Indonesia:ti,ab,kw OR Laos:ti,ab,kw OR Malaysia:ti,ab,kw OR "Mekong Valley":ti,ab,kw OR Myanmar:ti,ab,kw OR Philippines:ti,ab,kw OR Singapore:ti,ab,kw OR Thailand:ti,ab,kw OR Timor-Leste:ti,ab,kw OR Vietnam:ti,ab,kw OR "Solomon Islands":ti,ab,kw

AND

([mh Spine] OR Spine:ti,ab,kw OR spines:ti,ab,kw OR spinal:ti,ab,kw OR Vertebral:ti,ab,kw OR Vertebra:ti,ab,kw OR Vertebrae:ti,ab,kw OR thoracic:ti,ab,kw OR lumbar:ti,ab,kw OR sacrum:ti,ab,kw OR intervertebral:ti,ab,kw)

NOT ([mh animals] not [mh humans])

with Cochrane Library publication date from Jan 2000 to present
